# Untuned antiviral immunity in COVID-19 revealed by temporal type I/III interferon patterns and flu comparison

**DOI:** 10.1101/2020.08.21.20179291

**Authors:** Ioanna-Evdokia Galani, Nikoletta Rovina, Vicky Lampropoulou, Vasiliki Triantafyllia, Maria Manioudaki, Eleftherios Pavlos, Evangelia Koukaki, Paraskevi C. Fragkou, Vasiliki Panou, Vasiliki Rapti, Ourania Koltsida, Andreas Mentis, Nikolaos Koulouris, Sotirios Tsiodras, Antonia Koutsoukou, Evangelos Andreakos

**Author notes:** Corresponding author, e.mail. These authors contributed equally to this work (Ioanna-Evdokia Galani, Nikoletta Rovina).

## Abstract

A central paradigm of immunity is that interferon (IFN) mediated antiviral responses precede the pro-inflammatory ones, optimizing host protection and minimizing collateral damage^1,2^. Here, we report that for COVID-19 this does not apply. By investigating temporal IFN and inflammatory cytokine patterns in 32 COVID-19 patients hospitalized for pneumonia and longitudinally followed for the development of respiratory failure and death, we reveal that IFN-λ and type I IFN production is both diminished and delayed, induced only in a fraction of patients as they become critically ill. On the contrary, pro-inflammatory cytokines such as TNF, IL-6 and IL-8 are produced before IFNs, in all patients, and persist for a prolonged time. By comparison, in 16 flu patients hospitalized for pneumonia with similar clinicopathological characteristics to COVID-19 and 24 milder non-hospitalized flu patients IFN-λ and type I IFN are robustly induced, earlier, at higher levels and independently of disease severity, while pro-inflammatory cytokines are only acutely and transiently produced. Notably, higher IFN-λ levels in COVID-19 patients correlate with lower viral load in bronchial aspirates and faster viral clearance, and a higher IFN-λ:type I IFN ratio with improved outcome of critically ill patients. Moreover, altered cytokine patterns in COVID-19 patients correlate with longer hospitalization time and higher incidence of critical disease and mortality compared to flu. These data point to an untuned antiviral response in COVID-19 contributing to persistent viral presence, hyperinflammation and respiratory failure.

## Main

Coronavirus Disease 2019 (COVID-19), triggered by the betacoronavirus SARS-CoV-2, has become one of the worst pandemics of our time, causing high incidence of pneumonia, acute respiratory distress syndrome (ARDS) and death^3,4^. One of the most notable features of SARS-CoV2 infection is that it goes unnoticed for a remarkably prolonged period of time, running a course of a mild or uncomplicated illness for weeks until sudden and severe symptoms develop, in a subgroup of patients, requiring hospitalization, oxygen support and/or admission to an intensive care unit (ICU)^3,4^. This is consistent with an unusually long incubation period of the virus, ranging from 2 to 14 days, and an unusually long presence of it in the respiratory tract, often being detectable for over a month after initial infection by conventional molecular diagnostic tests^5,6^. By comparison, influenza virus infection, the main respiratory virus accounting for pneumonia hospitalizations till now, has an incubation time of 1 to 4 days, a short window of virus positivity of a few days, and an abrupt onset of symptoms causing pneumonia within 1–3 days^7,8^. Other frequent respiratory viruses such as respiratory syncytial viruses, rhinoviruses, parainfluenza viruses, metapneumonoviruses and regular human coronaviruses have also shorter incubation times (ranging from 1–5 days) and more rapid and acute manifestation of symptoms^9^, rendering SARS-CoV2 quite unique in that respect. The basis of this is unknown but is likely to be a key driver of the pathophysiology of COVID-19 underlying its distinctive disease course and clinical manifestations.

The hallmark of COVID-19 is the development of a hyper-inflammatory response, also known as ‘cytokine storm’, impairing the gas-exchange function and leading to acute respiratory distress syndrome (ARDS), multi-organ failure and death^10–12^. We and others have previously shown that a finely tuned antiviral response, orchestrated by IFN-λ (type III IFN) and type I IFN is critical for balancing immunity for optimal protection and minimal damage^13–15^. Deviation from this can unleash a detrimental ‘cytokine storm’ with devastating consequences for human health. A recent study suggested that in COVID-19 patients type I IFN and IFN-λ are not produced as they could not be detected in the sera of a small COVID-19 cohort of otherwise unspecified clinical characteristics^16^. In contrast, another one reported that type I IFN is induced in COVID-19 patients, and indicated that their levels might be reduced in those that are critically ill^17^. Such discrepancy could be due to the fact that each of these studies focuses on a single and likely distinct snapshot of an apparently heterogeneous disease process. Therefore, pursuing kinetic analyses is pertinent to delineating the course of the immune response, especially given that cytokines are transiently produced. This is particularly true for IFNs which are expressed early during infection and are rapidly down-regulated thereafter.

Here, we have performed a comprehensive temporal analysis of type I and type III IFN, and major inflammatory cytokine patterns in 32 COVID-19 and 16 influenza A virus infected (flu) patients hospitalized for community acquired pneumonia and longitudinally followed up according to current WHO guidelines^18^. Both groups of patients exhibited similar clinicopathological characteristics and comparable disease severity on admission (Table 1). We have also analyzed 24 milder flu patients with no radiological findings of pneumonia and no need for hospitalization (referred to as mild flu; Table 1), as well as 10 healthy individuals. Using high sensitivity Luminex and ELISA assays, we quantified 18 cytokines and chemokines relevant to antiviral immunity and hyperinflammation in patient sera collected at defined time intervals following hospital admission (Fig. 1a and S1aa). This aligns patients on the basis of the same clinical criteria of disease symptoms and severity, mainly the presence of pneumonia and the requirement for oxygen support.

**Table 1.** Clinical, laboratory and imaging findings of the study patients on hospital admission

| Characteristics | COVID-19 Patients | FLU Patients |  |
| --- | --- | --- | --- |
|  | Moderate-to-severe, hospitalized (N=32) | Moderate-to-severe, hospitalized (N=16) | Mild, non-hospitalized (N=24<sup>#</sup>) |
| **Age** |  |  |  |
| Median (IQR), yr | 63 (49-77) | 62.5 (52-73) | 46 (31-50) |
| Age ≥65 yrs, no (%) | 13 (41) | 8 (50) | 2 (8) |
| *Male, no (%)* | 22 (69) | 9 (56) | 13 (54) |
| **Smoking history, no (%)** |  |  |  |
| Never smoked | 22 (69) | 7 (44) | 8 (40) (N=20) |
| Former smoker | 4 (12) | 3 (19) | 1 (5) (N=20) |
| Current smoker | 7 (22) | 6 (37) | 11 (55) (N=20) |
| **Coexisting disorder, no (%)** |  |  |  |
| Any | 19 (59) | 13 (81) | 10 (50) (N=20) |
| Diabetes | 7 (22) | 2 (12.5) | 0 (0) (N=20) |
| Hypertension | 12 (37.5) | 8 (50) | 2 (10) (N=20) |
| Cardiovascular disease | 2 (6) | 2 (12.5) | 1 (5) (N=20) |
| COPD | 0 | 6 (37.5) | 2 (10) (N=20) |
| Asthma | 3 (9) | 1 (6) | 0 (0) (N=20) |
| Cancer/Hematological malignancy | 0 (0)/0 (0) | 1 (6)/0 (0) | 0 (0)/ 0 (0) (N=20) |
| Chronic renal disease | 0 (0) | 0 (0) | 0 (0) |
| Overweight/Obese | 17 (53)/8 (25) | N/A /2 (12.5) | N/A/ 0 (0) |
| Median interval from first symptoms on admission (IQR), days | 5 (2-7) | 1 (1-2.5) | 2 (1-2) (N=16) |
| **Fever on admission** |  |  |  |
| Patients, no/Total no (%) | 12 (37.5) | 16 (100) | 19 (100) (N=19) |
| Median temperature (IQR), °C | 37.5 (36.5-38.1) | 38.5 (38.4-39) | 38.6 (38.2-39) |
| **Symptoms on admission, no (%)** |  |  |  |
| Dyspnea | 21 (66) | N/A | N/A |
| Cough | 20 (62) | 11 (69) | 8 (40) (N=20) |
| Fatigue | 18 (56) | 12 (75) | 20 (100) (N=20) |
| Myalgia | 4 (12) | 8 (50) | 13 (65) (N=20) |
| Diarrhea/ vomiting | 6 (19) | 4 (25) | 4 (20) (N=20) |
| Median oxygen requirement (IQR, L/min) | 3 (2-6) | 9.5 (6-11) | 0 |
| CURB-65 score (IQR) | 1 (0-2) | 3 (1-3) | 0 |
| **Laboratory findings on admission** |  |  |  |
| Leukocytes (IQR) x 10<sup>9</sup>/L | 6.40 (4.61-8.52) | 7.27 (5.96-8.55) | 5.85 (4.26-7.39) (N=16) |
| Neutrophils (IQR) x 10<sup>9</sup>/L | 5.65 (3.32-7.45) | 5.08 (3.66-7.42) | 4.26 (2.48-5.13) (N=16) |
| Lymphocytes (IQR) x 10<sup>9</sup>/L | 0.95 (0.54-1.37) | 0.77 (0.59-1.48) | 1.07 (0.72-1.42) (N=16) |
| Monocytes (IQR) x 10<sup>9</sup>/L | 0.40 (0.28-0.57) | N/A | N/A |
| Platelets (IQR) x 10<sup>9</sup>/L | 190 (131-242) | 203 (180-242) | 200 (164-227) (N=16) |
| CRP mg/dl, (IQR) | 13.6 (3.64-19.4) | 5.5 (1.65-12.6) | 1.33 (0.82-2.40) (N=16) |
| Lactate dehydrogenase (LDH) (IQR), IU/L | 334 (203-455) | 300 (244-320) | 215 (200-250) (N=16) |
| Alanine aminotransferase (ALT) >40 IU/L | 29 (23-39) | 27 (20-48) | 16 (12-22.5) (N=16) |
| Ferritin (IQR), ng/ml | 677 (395-1237) | N/A | N/A |
| PCT (IQR), ng/ml | 0.08 (0.04 - 0.38) | N/A | 0.06 (0.06-0.08) (N=10) |
| **Chest X-ray findings on admission** |  |  |  |
| **Abnormal results, no (%)** | 30 (94)<sup>##</sup> | 15 (94)<sup>##</sup> | 0 (0) |
| Unilateral infiltrates | 4 (12.5) | 5 (31) | 0 (0) |
| Bilateral infiltrates | 26 (81) | 10 (63) | 0 (0) |

<sup>#</sup> In some cases, data available for fewer patients as indicated.
<sup>##</sup> X-ray negative patients were positive for unilateral or bilateral opacities on CT.
N/A: Not available

**Figure 1:**
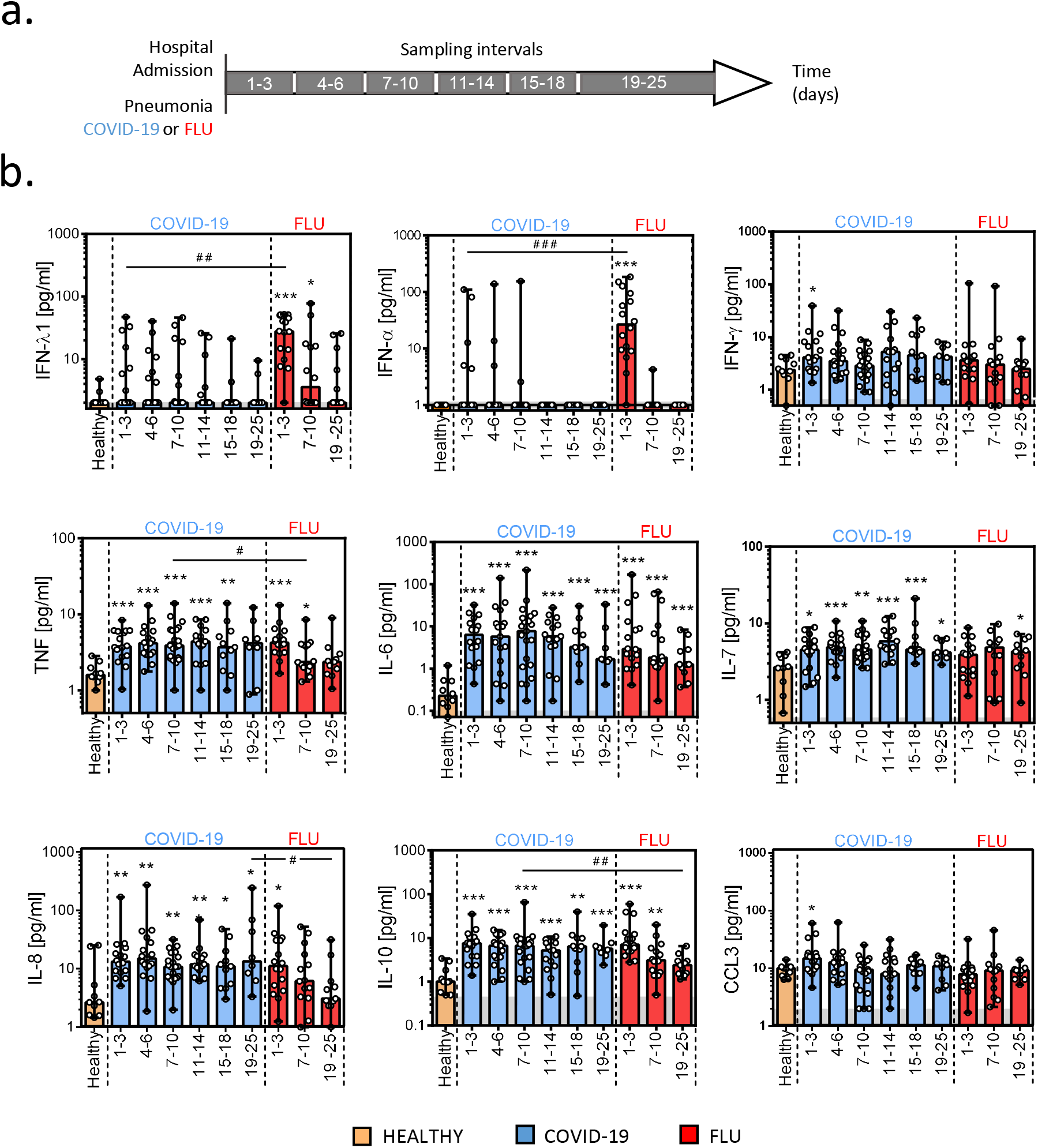
Temporal IFN and inflammatory cytokine patterns of COVID-19 and flu patients in relation to hospital admission. **a**, Schematic showing the experimental design with sampling at specific time intervals after hospital admission of 32 COVID-19 and 16 flu patients with pneumonia. **b**, Levels of IFN-λ1, IFN-α, IFN-γ, TNF, IL-6, IL-7, IL-8, IL-10 and CCL3 at various time intervals after hospital admission. Data are presented as scatter plots with dots showing individual patient measurements and columns median values with range. For COVID-19, n = 16, 17, 21, 15, 11 and 8 for each of the six consecutive time intervals. For flu, n = 16, 14 and 11, respectively. For healthy, n = 10. Grey shading marks the limit of quantification of the assay. *P* values were determined by a two tailed Mann–Whitney U test for nonparametric comparisons. \**P* < 0.05, \*\**P* < 0.01 and \*\*\**P* < 0.001 show significance over healthy controls.^#^*P* < 0.05,^##^*P* < 0.01 and^###^*P* < 0.001 show significance between COVID-19 and flu groups.

We found that COVID-19 patients had profoundly impaired induction of both IFN-λ and type I IFNs. Median levels of IFN-λ and type I IFNs were not detectable in most COVID-19 patients (Fig. 1b), although some patients made IFN-λ and fewer of them also IFN-α. This contrasts flu patients who almost uniformly expressed both types of IFNs, within the first (day 1–3) time interval of admission, and at significantly higher levels. At all cases, IFN expression was transient, with type I IFN levels rapidly declining after the first three days of hospitalization, while IFN-λ persisting longer. Interestingly, despite their limited ability to make IFNs, COVID-19 patients robustly expressed pro-inflammatory cytokines such as TNF, IL-6, IL-7, IL-8, IL-10, IFN-γ and CCL3 that were maintained at high levels for a prolonged time (Fig. 1b). Other cytokines such as IL-1β, IL-12, IL-23 and CCL4 were also significantly up-regulated at specific time intervals compared to healthy individuals reflecting the heterogeneity of the disease course (Fig. S2).

A similar pattern emerged when comparisons were made according to disease symptoms onset (Fig. S1b). COVID-19 patients exhibited markedly delayed and reduced IFN-λ and type I IFN levels which were detectable only in a fraction of the patients and from days 7–10 onwards of symptoms onset (Fig. S3, a-b). By comparison, all flu patients exhibited high levels during the first 6 days (Fig. S3, a-b). Although COVID-19 patients made little IFN during the first 6 days of symptom onset, they potently produced pro-inflammatory cytokines and chemokines such as TNF, IL-6, IL-8, IL-10 and CCL3 at levels similar to flu (Fig. S3, b-c). Moreover, they exhibited prolonged expression of pro-inflammatory mediators, with high levels of TNF, IL-6, IL-7, IL-8, IL-10 and CCL4 remaining detectable for over three weeks of onset, whereas in flu patients a number of these were by that time down-regulated (Fig. S3).

Notably, COVID-19 patients were admitted to hospital with similar markers of systemic inflammation such as CRP levels, white blood cell (WBC) and neutrophil counts, and neutrophil/lymphocyte (N/L) ratio to flu patients (Table 1 and Fig. S4, a-f). They even had lower fever and a lower CURB-65 score, a commonly used measure of pneumonia severity^19^ (Fig. S4, g-h). However, during follow up COVID-19 patients developed a much higher incidence of ARDS requiring ICU support. In our cohort, 16 out of 32 patients (50%) developed critical disease, 3 of which died, compared to only 3 out of 16 flu patients(18.7%) none of which died (Fig. 2, a-b). Strikingly, COVID-19 patients became critically ill over a much broader time window (up to nine days after admission) than flu patients which manifested critical disease within the first day post admission. This is in agreement with the high incidence and protracted course of severe respiratory failure described for COVID-19^4,12^. Interestingly, among COVID-19 patients those who became critically ill had higher CRP levels, WBC and neutrophil counts, and N/L ratio on admission (Fig. S4, a-f), but not CURB-65 or fever (Fig. S4, g-h and Table S1). Critically ill flu patients also had a tendency for higher WBC and neutrophil counts and a N/L ratio, as well as significantly raised CURB-65, whereas non-hospitalized flu patients did not exhibit any of these increases (Fig S4, ah).

**Figure 2:**
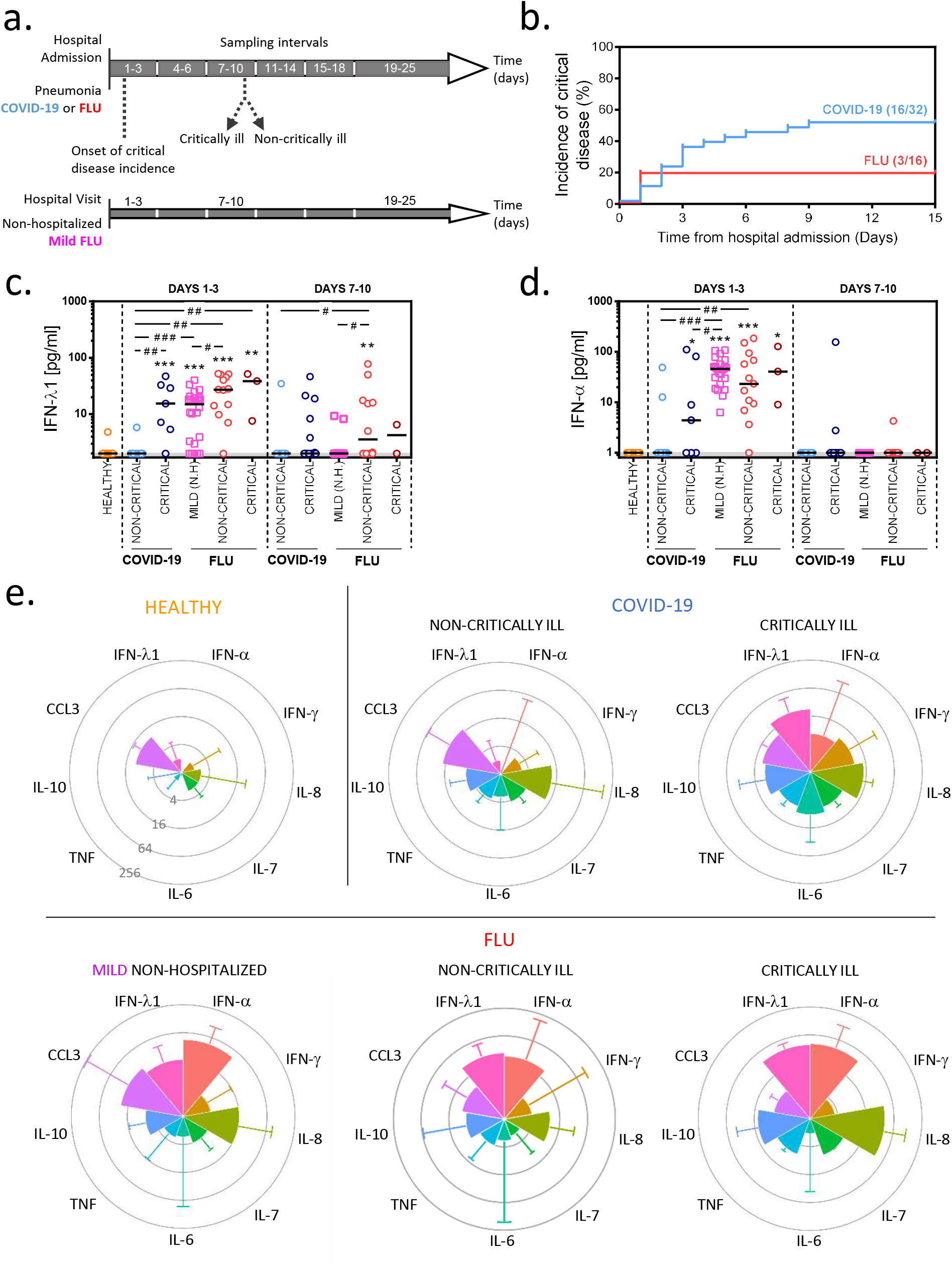
Comparison of IFN and inflammatory cytokine patterns between subgroups of COVID-19 and flu patients according to disease severity. **a**, Schematic depicting the longitudinal follow up of 32 COVID-19 and 16 flu patients hospitalized for pneumonia, and 24 non-hospitalized (N.H.) flu patients with no radiological findings of pneumonia. **b**, Incidence of critical disease in hospitalized COVID-19 and flu patients over time. **c-d**, Levels of IFN-λ1 (c) and IFN-α (d) of critically and non-critically ill patients, and mild non-hospitalized patients at day 1–3 and 7–10 time intervals after hospital admission or visit, respectively, as well as healthy individuals. Dots show individual patient measurements and lines median values of hospitalized patients and healthy individuals. Squares show non-hospitalized flu patients. Grey shading marks the limit of quantification of the assay. **e**, Radar plots of median cytokine levels and range of hospitalized COVID-19 and flu patients developing critical versus non-critical illness, non-hospitalized flu patients and healthy individuals at the day 1–3 time interval after admission. Each circle in the radar plot represents logarithmically increasing concentrations from 4 to 256 pg/ml as shown in the healthy control. For days 1–3 n = 9, 7, 24, 13 and 3 for each of the five consecutive groups, respectively. For days 7–10 n = 8, 13, 15, 12 and 2, respectively. For healthy individuals, n = 10. *P* values were determined by a two tailed Mann–Whitney U test for non-parametric comparisons. \**P* < 0.05, \*\**P* < 0.01 and \*\*\**P* < 0.001 show significance over healthy controls.^#^*P* < 0.05,^##^*P* < 0.01 and^###^*P* < 0.001 show significance between COVID-19 and flu subgroups.

We thus examined whether temporal cytokine patterns differ between the various patient groups. Surprisingly, we observed that although COVID-19 patients that do not become critically ill produce little type I or III IFN, the ones that become critically ill make IFN-λ which are significantly higher at the day 1–3 time interval compared to healthy and non-critically ill patients (Fig. 2c and S5). Some of the critically ill patients also make IFN-α (Fig. 2d and S5), albeit at significantly lower levels to mild non-hospitalized flu patients (Fig. 2d) or the total of hospitalized flu patients (both critically and non-critically ill; p< 0.05). On the contrary, all COVID-19 patients make pro-inflammatory cytokines such as TNF, IL-6, IL-8, IL-10 and IFN-γ, with critically ill patients exhibiting also higher levels of IL-6, IL-7 and TNF at specific time intervals, and a tendency for higher IFN-γ consistent with the increased hyper-inflammatory state they are in (Fig. 2e, S5 and S6). Individual patient data further confirmed these trends (Fig. S7). Interestingly, CCL3 is significantly higher than healthy controls in non-critically ill COVID-19 patients but not in the critically ill ones (Fig. S6). By comparison, critically ill and non-critically ill flu patients did not differ in their ability to make type I and type III IFNs nor pro-inflammatory cytokines such as TNF, IL-6 or IL-7 (Fig. 2, c-e, S5 and S6). Similarly, non-hospitalized flu patients with mild disease exhibited strong production of type I and type III IFNs, indicating that across the spectrum of flu disease severity the antiviral response remains robust. Visualizing these patterns on radar plot reveals a major imbalance in the induction of antiviral and pro-inflammatory responses of COVID-19 patients that does not occur in flu (Fig. 2e).

We next sought to determine whether imbalanced cytokine patterns in COVID-19 patients are related to systemic immune effects, and parameters linked to disease severity. To that end, we obtained temporal white blood cell transcriptomes from 5 healthy individuals and 9 COVID-19 patients, 5 non-critically and 4 critically ill, starting from day 1 of entry to the ward or ICU and at different timepoints thereafter. In total, 24 comprehensive RNAseq gene expression datasets were analyzed, clustering according to the clinical phenotype and indicating this as the main source of variation (Fig. 3a and S8a). Focusing at day 1 as the most relevant timepoint, we found that 4225 genes were differentially expressed (DEGs) in COVID-19 patients compared to healthy individuals (Table S2). When critically and non-critically ill patients were compared separately to healthy controls, 4225 and 4902 DEGs were observed, respectively, of which 1979 were common whereas the rest were uniquely found in one or the other patient group (Fig. S8b, Table S2 and Table S3). Volcano plots pointed out notable differences in the most highly regulated genes between the groups with critically ill patients exhibiting a stronger immune and antiviral response gene patterns (Fig. S8, c-e). Pathway analysis of DEGs indeed revealed that the most significant pathways over-represented in critically ill patients were related to the positive regulation of the immune system, the activation of the innate immune response, the defense response to virus and the cellular response to IFN (Fig. 3b and Table S4). By contrast, in non-critically ill patients these pathways were not significant; the ones over-represented instead included the regulation of the cellular component size, IL-1β production and NK cell cytotoxicity (Fig. 3b).

**Figure 3:**
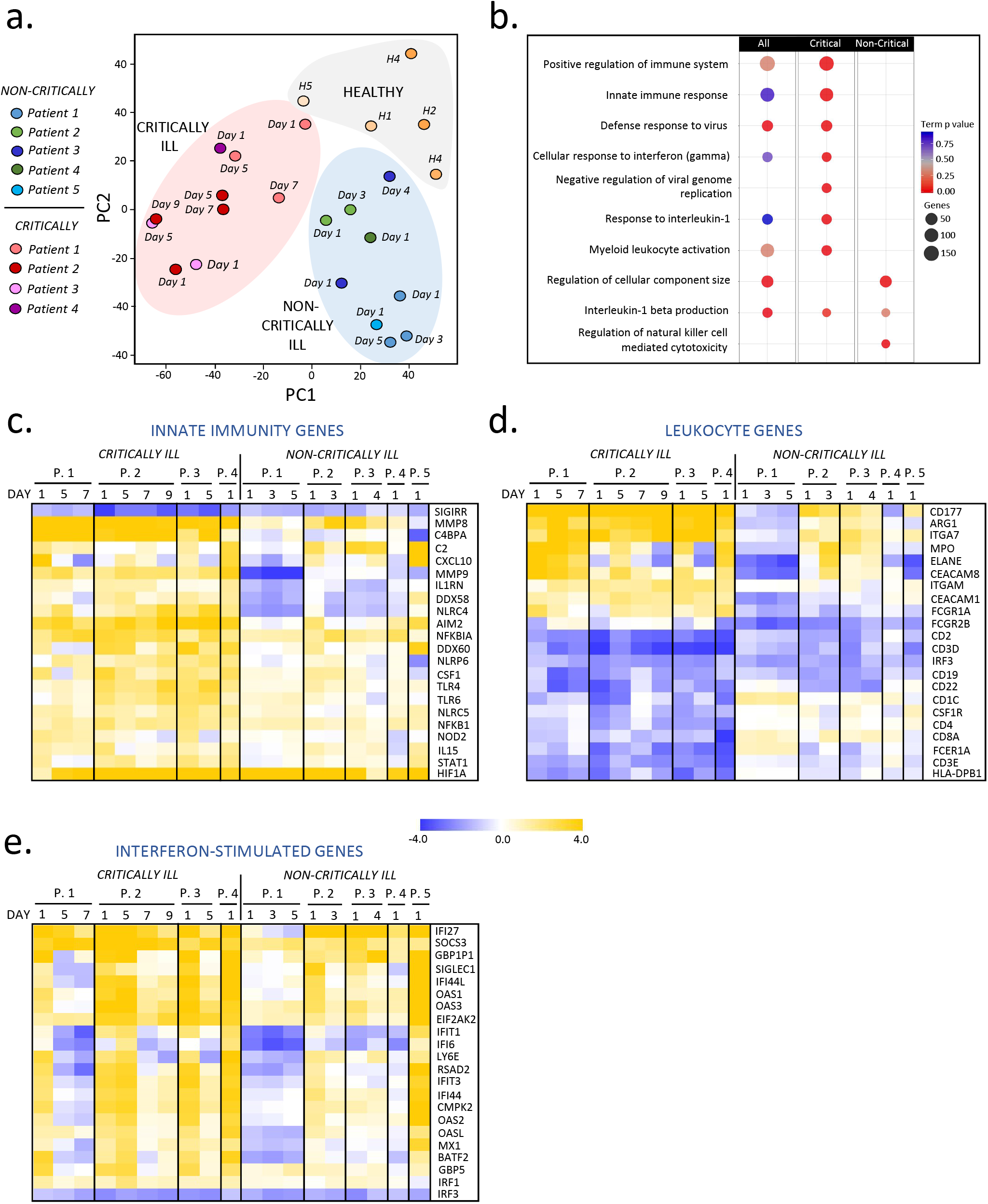
Kinetic analysis of blood transcriptional signatures of critically and non-critically ill COVID-19 patients. **a**, Principal component analysis of peripheral white blood cell transcriptomes of critically ill (n=4) and non-critically ill (n=5) patients or healthy (H1-H5) controls (n=5). **b**, Gene ontology (GO) pathway enrichment analysis of differentially expressed genes of all, critically ill and non-critically ill patients at day 1 of hospital or ICU entry. **c-e**, Heatmaps of differentially expressed innate immunity, leukocyte and IFN-stimulated genes and their kinetics following hospitalization of critically ill and non-critically ill COVID-19 patients compared to healthy controls. Data are expressed as log2 fold values over healthy controls.

Accordingly, heatmaps with temporal information unveiled the systemic activation of the innate immune response marked by the up-regulation of key pro-inflammatory genes and pattern recognition receptors (*C4bpa, Csf1, Il1rn, Cxcl10, Mmp8, Stat1, Ddx58/Rigi, Tlr4, Nlrp6*), and the induction of a dominant activated neutrophil-myeloid cell signature (*Mpo, Elane, Cd177, Itgam, Arg1, Ceacam8, Fcgr1a*) in the critically ill group that was milder and not significant in non-critically ill patients (Fig. 3, c-d). On the contrary, T and B lymphocyte lineage and related genes (*Cd3d, Cd3e, Cd4, Cd8a, Cd19, Cd22*) were markedly down-regulated in critically ill patients. These data are consistent with lymphopenia, high neutrophil counts, and a high N/L ratio also present in these patients (Fig. S4) and previously reported to be associated with more severe disease and worse outcomes in COVID-19 patients^3,4^. Cytokines such as TNF, IL-6 and IL-8 may directly account for these effects, as they are well known to trigger the mobilization and activation of neutrophils, the development of lymphopenia and the induction of innate immune responses and systemic inflammation^20,21^. Notably, a long set of IFN-stimulated genes (ISGs) was also strongly induced in critically ill patients compared to only a fraction of them being up-regulated in the non-critically ill group (Fig 3e and S8f), in agreement with the patterns of IFN-λ and type I IFN production in these patients. This cannot be attributed to differential expression of IFN receptors as no differences between Ifnlr1, Il10rb, Ifnar1 and Ifnar2 mRNA levels were observed among patient groups and healthy individuals with the exception of a 2-fold up-regulation of the already high levels of Ifnar2 in critically ill patients (Table S2).

Interestingly, imbalanced cytokine patterns in COVID-19 patients with pneumonia were associated with a much worse disease outcome compared to flu. First, the COVID-19 group exhibited higher incidence of critical disease and mortality (Fig. 2b). Second, COVID-19 patients overall, as well as when grouped as critically and non-critically ill, required longer hospitalization time than their flu counterparts (Fig. 4, a-c). For non-critically and critically ill COVID-19 patients, median time was 14 and 23 days, respectively, compared to flu that was 7 and 19 days (Fig. 4, b-c). Prolonged hospitalization could be attributed to the untuned antiviral responses, leading to a more protracted clinical course of COVID-19 relative to flu and a need for longer recovery even for the non-critically ill group. To identify cytokines and cytokine combinations that can predict hospitalization time and therefore be of prognostic value, we generated a correlation matrix of the cytokine levels at admission (days 1–3 interval) and the duration of hospital stay (Fig. 4d). We found that higher IL-6 and IL-10, and lower CCL3 levels, are directly proportional to the duration of hospitalization (Fig. 4, d-f). The value of TNF and IL-6 as biomarkers for monitoring COVID-19 severity has been reported^4,22^,^23^ but for CCL3 this is new. Interestingly, IFN-λ levels also correlated with higher IL-6, and longer hospitalization, consistent with their almost exclusive induction in critically but not non-critically ill patients (Fig. 4d and 2c).

**Figure 4:**
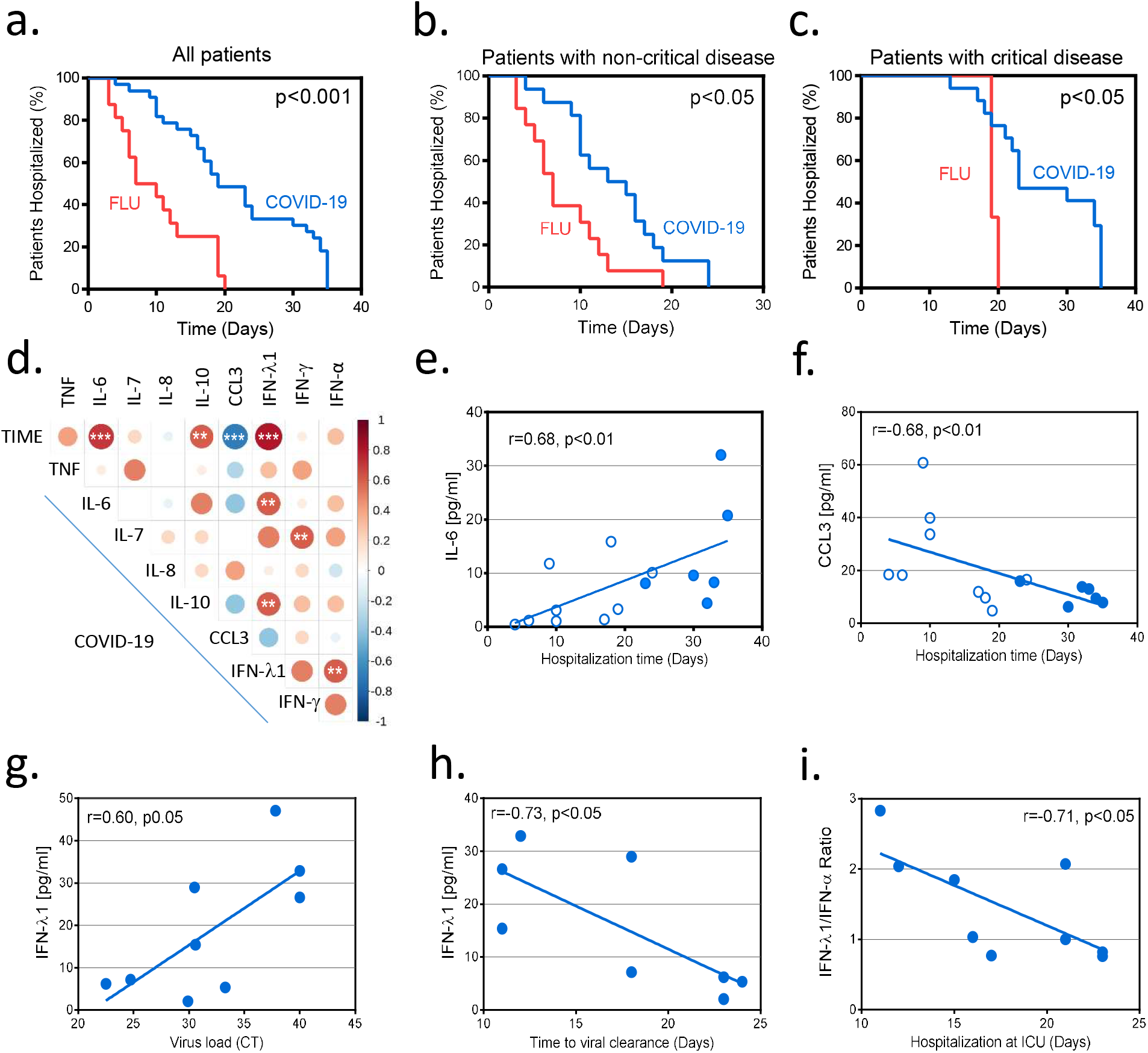
Correlation of IFN and cytokine expression patterns with disease outcomes. **a-c**, Comparison of hospitalization time between COVID-19 and flu patients. All patients (a), patients with non-critical disease (b) and patients with critical disease (c) are shown. **d**, Correlation matrix of cytokine concentration levels at the day 1–3 time interval after hospital admission of COVID-19 patients indicating correlations between cytokines and total hospitalization time (TIME) or other cytokines. **e-f**, Correlation of IL-6 (e) and CCL3 (f) concentration levels with the duration of total hospitalization of all COVID-19 patients. **g**, Correlation of IFN-λ1 concentration levels with viral load expressed as CT values in bronchial aspirates collected at the same time interval as the sera used for IFN-λ1 quantification. **h**, Correlation of IFN-λ1 concentration levels with time required for viral clearance assessed as the first – ve SARS-CoV2 test. **i**, Correlation of IFN-λ1:IFN-α ratio with the duration of hospitalization in the ICU. Dots show individual COVID-19 patient measurements and shaded dot plots values corresponding to COVID-19 patients becoming critically ill. *P* values were determined using the Spearman rank-order correlation coefficient for non-parametric data. \**P* < 0.05, \*\**P* < 0.01 and \*\*\**P* < 0.001

A question that arises is whether IFN levels induced in critically ill patients are beneficial as delayed type I or type III IFN production has been shown in animal models to cause immunopathology^13,14^,^24^ or interfere with epithelial repair^25,26^, respectively. We found that higher IFN-λ concentrations during ICU entry were associated with lower SARS-CoV2 viral load in the respiratory tract and faster viral clearance (Fig. 4, g-h). Moreover, a higher IFN-λ to type I IFN ratio at that time was linked to a shorter stay in the ICU (Fig. 4i), with the two patients with the highest IFN-α levels also exhibiting the longest stay (both 23 days over a median of 17 days). These data suggest that delayed IFN-λ induction may still be protective in critically ill COVID-19 patients whereas IFN-α may do more harm than good, at least in a subset of patients.

Taken together, our findings demonstrate that SARS-CoV2 infection does not follow the conventional paradigm of antiviral immunity. Instead of activating first the antiviral response followed by the pro-inflammatory process as a second line of protection, it does the opposite; it triggers the pro-inflammatory response long before IFN-mediated antiviral defenses are induced- if at all. This is a major paradox and helps explain many of the unique or unusual features of COVID-19. The long virus incubation time and persistence in the respiratory tract, giving positive SARS-CoV2 tests for weeks, can be attributed to the delayed and/or reduced production of type I and III IFNs. The absent or very mild symptoms of patients for an unusually extended period of time, can be attributed to the lack or impaired and delayed expression of type I IFNs, principal mediators of flu-like disease and symptoms such as runny nose, coughing, fatigue, dyspnea and fever in humans^27^. Finally, the early and persistent expression of pro-inflammatory cytokines culminating into prolonged hyper-inflammation can promote the sudden development of respiratory failure requiring hospitalization and frequently ICU admission. Noteworthy, in flu the swift induction of the type I and III IFN response, across the spectrum of disease severity, correlates with quicker recovery, and markedly lower incidence of critical disease or mortality^13,24^. The recent demonstration in a retrospective cohort study of 446 COVID-19 patients that early administration of IFN-α (interferon-a2b) is linked to reduced in-hospital mortality whereas late IFN-α therapy leads to increased mortality and delayed recovery leaves little doubt that the timing of IFN production is also crucial in COVID-19 patients^28^. Conceivably, late production of type I or III IFN production might confer no viral resistance, but instead promote immunopathology.

Whether this unique clinical course of COVID-19 is related to the presence of SARS-CoV2-derived IFN inhibitors as previously proposed for SARS-CoV^29,30^ and MERS-CoV^31^ is not known but is a possibility. As with other viruses, inhibition may be overcome once higher viral loads are reached, e.g. after incubation of the virus and eventual spread in susceptible individuals. In our study, we did not see significant differences in virus levels between non-critically and critically ill patients at the time IFNs were measured (Fig. S9). However, higher virus load in severe over mild disease has been described in one study but not been confirmed in another^32,33^. Moreover, higher virus load can overcome SARS-CoV2 dose-dependent suppression of IFN production in cultured respiratory epithelial cells^16^.

Our study is not without caveats. First, it characterizes cytokine patterns in the circulation, and although these are commonly used to analyze ‘cytokine storms’ in response to infection, how well they correlate to immune responses in the respiratory tract is difficult to know. Second, it is relatively small, and our findings await validation in other cohorts. Still, our study is uniquely informative as it addresses the production of IFNs and the activation of the ‘cytokine storm’ in COVID-19 in a temporal manner, from hospital admission to ICU entry, and should therefore be particularly useful for the design of clinical trials testing IFN therapies. Finally, it provides a side-by-side comparison of COVID-19 with flu, studying patient populations with similar genetic, demographic and clinicopathological characteristics, and therefore uncovers important differences in the antiviral immune response between these two diseases that have not been previously suspected.

## Methods

### Study participants

In this non-interventional study thirty two patients with diagnosis of COVID-19 pneumonia according to WHO guidelines and positive SARS-CoV-2 reverse transcription polymerase chain reaction (RT-PCR) testing on a respiratory sample (nasopharyngeal swab or bronchial aspirate) were included^18^. Patients were recruited between March and April 2020 from the 1^st^ Respiratory and Critical Care Clinic ward and ICU of the “Sotiria” General Chest Diseases Hospitalof Athens, Greece. Healthy, asymptomatic subjects with a negative SARS-CoV-2 RT-PCR at the time of inclusion served as the control group.

The severity of COVID-19 cases was classified based on the adaptation of the Seventh Revised Trial Version of the Novel Coronavirus Pneumonia Diagnosis and Treatment Guidance^34^. All patients had moderate to severe disease, and presented with respiratory symptoms and radiological findings of pneumonia. They met any of the following criteria:

1. Respiratory distress (≥30 breaths/ min);
2. Oxygen saturation ≤93% at rest;
3. Arterial partial pressure of oxygen (PaO2)/ fraction of inspired oxygen (FiO2) ≤300mmHg with no other organ failure.

Sixteen patients developed ARDS and critical illness due to respiratory failure (PaO2/FiO2 ≤ 200 mmHg) requiring mechanical ventilation, with shock and/or other organ failure necessitating intensive care unit (ICU) care.

Blood was drawn at various time intervals during hospitalization and at discharge; and white blood cells and plasma were collected and stored. Serum was also stored, almost daily, for further use. In order to better understand the immune response of COVID-19 infection, subjects from a cohort of patients with confirmed H1N1/H3N2 influenza A virus (flu) infection were also studied. In total, 40 patients were recruited at the 2^nd^ Respiratory Clinic of the ‘Sotiria’ General Chest Diseases Hospital, Athens, Greece and the “Attikon” University Hospital, University of Athens Medical School, Athens, Greece. Confirmation was obtained from nasopharyngeal swabs using the BioFire FilmArray Respiratory Panel test (bioMerieux). Patients were categorized according to the severity of the disease into mild flu patients with no radiological findings of pneumonia, no need for oxygen support and hospitalization, and moderate to severe flu patients with radiological findings of pneumonia (x-ray or CT), oxygen need and symptoms requiring hospitalization. Hospitalized flu patients could be subdivided into patients that did not develop (PaO2/FiO2 > 200 mmHg) or developed critical disease (PaO2/FiO2 ≤ 200 mmHg). Flu patients had similar clinico-pathological characteristics to COVID-19 patients upon admission (Table 1). All subjects included in the study were clinically evaluated and followed longitudinally during the whole period of hospitalization (from admission to discharge). All blood specimens were processed immediately for serum collection and aliquots were stored at –80^°^C.

The study conforms to the principles outlined in the Declaration of Helsinki, and received approval by the Ethics Committees of the “Sotiria” General Chest Diseases Hospital, Athens, Greece (Approval numbers 16707/10–7–18 and 8385/31–3–20) and the “Attikon” University Hospital, University of Athens Medical School, Athens, Greece (Approval number 1821A/22–9–16).

### SARS-CoV-2 detection

RNA was extracted from nasopharyngeal swabs and bronchial aspirates by using the Nuclisens easyMAG instrument (bioMerieux), according to the manufacturer’s instructions. Real time RT-PCR was performed on extracted nucleic acids targeting the E gene of SARS-CoV-2 as described by Corman et al^35^.

### Cytokine analysis

Serum samples frozen and stored at −20°C, without other thawing, were analyzed for the presence of IFNγ, TNF, IL-1β, IL-2, IL-4, IL-6, IL-7, IL-8, IL-10, IL-12 (p70), IL-13, IL-17A, IL-23, CCL3, CCL4 and CX3CL1 with the MILLIPLEX MAP Human High Sensitivity T cell Panel (Merck Millipore). Thawed serum aliquots were centrifuged at 13,000 rpm for 10 min at 4°C immediately prior to testing. Each assay was performed according to the manufacturer’s protocol for serum samples, utilizing recommended sample dilutions and standard curve concentrations (Merck Millipore). Samples were analyzed on a Luminex 200™ System according to the manufacturer’s instructions (Merck Millipore). For each cytokine on each assay, the lowest detection limits were in pg/ml: 0.50 for IFN-γ, 0.42 for TNF, 0.2 for IL-1β, 0.24 for IL-2, 0.60 for IL-4, 0.16 for IL-6, 0.33 for IL-7, 0.30 for IL-8, 0.50 for IL-10, 0.24 for IL-12 (p70), 0.20 for IL-13, 0.50 for IL-17A, 8.00 for IL-23, 2.00 for CCL3, 0.80 for CCL4 and 10.00 for CX3CL1. High sensitivity sandwich Enzyme-linked Immunosorbent Assay (ELISA) kits were used for the detection of human IFN-α (Thermo Scientific) and IFN-λ1 (Biolegend). Their sensitivity in pg/ml was 1.00 for IFN-α and 2.00 for IFN-λ1.

### RNAseq analysis

For RNAseq analysis, total RNA was purified from whole blood leukocytes with the RNeasy Micro kit (Qiagen). RNA samples were treated with DNase I (Qiagen) and quantified on a NanoDrop (Thermo Scientific). NGS libraries were prepared with the TruSeq RNA Library Prep Kit v2 (Illumina) according to the manufacturer’s instructions. Quality of the libraries was validated with an Agilent DNA 1000 kit run on an Agilent 2100 Bioanalyzer. Library samples were quantified using a Qubit^™^ dsDNA HS Assay Kit (Thermo Scientific) according to the manufacturer’s instructions. Bar-coded cDNA libraries were pooled together in equal concentrations in one pool, and were sequenced on a NextSeq 500 System (Illumina) at the Greek Genome Center (Biomedical Research Foundation, Academy of Athens, Athens, Greece).

### Transcriptomics analyses

Samples sequenced on NextSeq 500 (Illumina) were analyzed using standard protocols. Briefly, raw reads were pre-processed using FastQC v.0.11.2 and cutadapt v.1.6, and then mapped to the human genome (GRCh38) using the TopHat version 2.0.13, Bowtie v.1.1.1 and Samtools version v.1.1. The read count table was produced using HTSeq v.0.6. Following filtering of raw read counts with a threshold of 10 in at least one dataset, resulting in a total of 21880 genes, DESeq2 analysis was performed^36^. This returned the log2foldchanges of the treatment compared to healthy individuals for each time point. Differentially expressed gene transcripts were selected based on an adjusted *p*-value cutoff of 0.05 (FDR 5%). Pathway enrichment analysis was conducted using ClueGO and CluePedia plugin of Cytoscape. Heatmaps were performed using TM4 MeV v.4.8 and Euclidean distance was used for hierarchical clustering. Clustering and dendrograms were performed with hclust function and ggdendro package, respectively, in R.

### Data and software availability

The raw RNAseq data have been deposited at GEO (http://www.ncbi.nlm.nih.gov/geo/) under BioProject accession number # PRJNA638753.

### Statistical analysis

Data were analyzed on GraphPad Prism software. Statistical significance of differences was assessed using the Mann-Whitney *U* (MWW) test for non-parametric data. Associations between cytokines and hospitalization time (in days) were tested using Spearman rank-order correlation coefficient and visualized using the corrplot R package. Polar charts from the ggplot2 R package were used for the visualization of the differences in cytokine response patterns.

## Acknowledgements

The authors would like to thank Dr A. Gavriil and G. Vatsellas for technical assistance, and Prof. P. Katsikis and V. Soumelis for critically reading the manuscript. EA is supported by research grants from the European Commission (IMMUNAID, No 779295 and CURE, No. 767015), the Hellenic Foundation for Research and Innovation (INTERFLU, No 1574) and Janssen Pharmaceuticals. IEG is supported by a research grant from the Hellenic Foundation for Research and Innovation (RELIEVE, No 506). This work received also a donation from the J. Sanchez family and friends.

## Conflict of interest

The authors declare that they have no conflict of interest.

## Notes

### Competing Interest Statement

The authors have declared no competing interest.

### Author Declarations

The study conforms to the principles outlined in the Declaration of Helsinki, and received approval by the Ethics Committees of the Sotiria General Chest Diseases Hospital, Athens, Greece (Approval numbers 16707/10-7-18 and 8385/31-3-20) and the Attikon University Hospital, University of Athens Medical School, Athens, Greece (Approval number 1821A/22-9-16).

